# Polygenic Risk Score Modifies Risk of Coronary Artery Disease Conferred by Low-Density Lipoprotein Cholesterol

**DOI:** 10.1101/2020.03.01.20029454

**Authors:** Alessandro Bolli, Paolo Di Domenico, Roberta Pastorino, George Busby, Giordano Bottà

## Abstract

**Background:** An individual’s lifetime risk of Coronary Artery Disease (CAD) is determined by a combination of genetic and lifestyle factors. Whilst adherence to a healthy lifestyle can help individuals with high genetic risk reduce their lifetime risk of CAD, the extent to which blood lipid levels affect CAD risk in individuals with varying genetic risk remains unknown. To explore how genetics, blood lipids and CAD risk interact, we derived a novel genome-wide polygenic risk score (PRS) for CAD. We then applied the PRS to individuals from the UK Biobank and divided them into Low PRS (bottom 10 percentiles of PRS distribution), Intermediate PRS (PRS in the 10th-90th percentiles), and High PRS (top 10 percentiles), and further stratified individuals by blood lipid levels.

**Results:** We found that the elevated CAD risk conferred by high low-density lipoprotein cholesterol (LDL-C) was modified by the interaction with PRS (P-value interaction: <0.005). Individuals with High PRS and whose LDL-C was Borderline (between 130 and 160 mg/dL) had higher CAD relative risk (HR 3.10; 95% CI, 2.55-3.76) than those at Intermediate PRS whose LDL-C were Very High (>190 mg/dL; HR 2.77; 95% CI, 2.33-3.28). Furthermore, individuals with High PRS but whose lipid levels were below the following thresholds did not have a significantly increased risk for incident CAD: LDL-C <130 mg/dL, total Cholesterol (TC) <200 mg/dL, LDL-C:HDL <2.0 and TC:HDL <3.0. In addition, individuals with Low PRS and Very High LDL-C (>190 mg/dl) did not have increased CAD risk, which was comparable to individuals with Intermediate PRS and Optimal LDL-C (<130 mg/dL).

**Conclusions:** Our results have important implications for the primary prevention of coronary artery disease. Currently, healthy individuals with Borderline LDL-C (130-159 mg/dL) are not considered to be at high risk of CAD. Here we demonstrate that the combination of Borderline LDL-C and High PRS results in CAD relative risk which is greater than individuals without high polygenic risk, but whose LDL-C levels are high enough for statins to be recommended (>190 mg/dL). This analysis therefore demonstrates that PRS can identify a proportion of the population who are at high-risk of CAD but who are invisible to current approaches for assessing CAD risk. Moreover, of perhaps greater significance is the evidence that individuals who have a combination of High PRS and Optimal blood lipid levels do not have greater risk of CAD than individuals without high polygenic risk and the same Optimal blood lipid levels. Our results suggest that high polygenic risk for CAD could be overcome by controlling blood lipid levels. We propose that incorporating PRS into CAD risk assessment early in life could allow individuals at high polygenic risk to benefit from tailored blood lipid guidelines and avoid lifetime exposure to potentially damaging PRS-dependent LDL-C levels.

## Introduction

Coronary artery disease (CAD) is the leading cause of death worldwide (Lozano et al., 2012). CAD is a complex disease and its etiology is characterised by the complicated interplay between both genetic and lifestyle factors. 2,213 genetic variants have been found to be significantly associated with CAD (Nikpay et al., 2015), highlighting the polygenic architecture of the disease. Effect sizes of risk-associated alleles identified in Genome-Wide Association Studies (GWAS) can be combined to generate a Polygenic Risk Score (PRS), a quantitative metric used to stratify individuals at different polygenic risks (Torkamani et al., 2018). A CAD PRS developed by Khera et al. (2018) identified that 8% of individuals of the UK Biobank have an increased risk of CAD that is more than three times higher than the remainder of the population, which is similar to the risk conferred by familial hypercholesterolemia. In a second study, men in the top 20% of a CAD PRS distribution reached a threshold of 10% cumulative CAD risk by 61 years of age, more than 15 years earlier than men in the bottom 20% of the distribution (Inouye et al., 2018).

CAD PRS can be used in the general population to identify those individuals that would benefit the most from targeted lifestyle recommendations and preventive interventions. A high genetic risk of CAD has been shown to be actionable in two ways. First, individuals at high genetic risk of CAD who adhere to a healthy lifestyle display a two- (Khera et al., 2016) or three-fold (Said et al., 2018) reduction in the relative risk of developing CAD, compared to individuals with a poor lifestyle. Second, in a randomized controlled trial of men with hypercholesterolemia treatment with lipid-lowering medications reduced CAD risk from 19.6% to 11.7%, in individuals in the top quintile of the PRS distribution, and from 12.9% to 10.1%, in the remaining individuals despite both groups had same LDL level at baseline (LDL-C 192 mg/dL), and achieved almost identical LDL levels after treatment (148 and 149 mg/dL, High PRS and remainder, respectively). This finding suggests that the polygenic background may act as an enhancer, increasing CAD risk by facilitating the LDL-C-dependent process of plaque formation (Ference et al., 2018).

Despite such evidence, a systematic assessment of the interplay between polygenic risk and blood lipids in contributing to the overall CAD risk is still lacking. To address this, we ask the following questions: **i)** How does the PRS odds ratio range compare to those of blood lipids commonly used to estimate CAD risk? **ii)** Is PRS correlated to conventional risk factors? **iii)** Is CAD risk conferred by blood lipids modulated by the polygenic background? **iv)** If and to what extent optimal blood lipid levels are associated with reduced CAD risk in individuals with High PRS?

## Methods

### Data sources

This study utilises the UK Biobank population (project number: 40692). Full details of the UK Biobank resource have been described previously (Bycroft et al., 2018). Briefly, between 2006 and 2010, the UK Biobank recruited around 500,000 participants between the ages of 40 and 70 years. At recruitment, information was collected for each participant including physical and functional measurements, demographic characteristics, and self-reported prevalent medical conditions. Incident events throughout the follow-up were recorded and accessed through electronic health care records. Additionally, a wide range of biochemical markers from blood and urine samples was collected at baseline, including risk factors such as blood lipid levels and associated (apo)lipoproteins. Each UK Biobank individual has been genotyped at ∼850,000 Single Nucleotide Polymorphisms (SNPs) which have subsequently been imputed to over 93M SNPs.

We selected an initial sub-population comprising 408,188 individuals of white British ancestry. Exclusion criteria comprised the mismatch between genetic and reported sex, outliers in heterozygosity and missing rates, Sex chromosome aneuploidy, and consent withdrawal (**Figure 1**).

**Figure 1.**
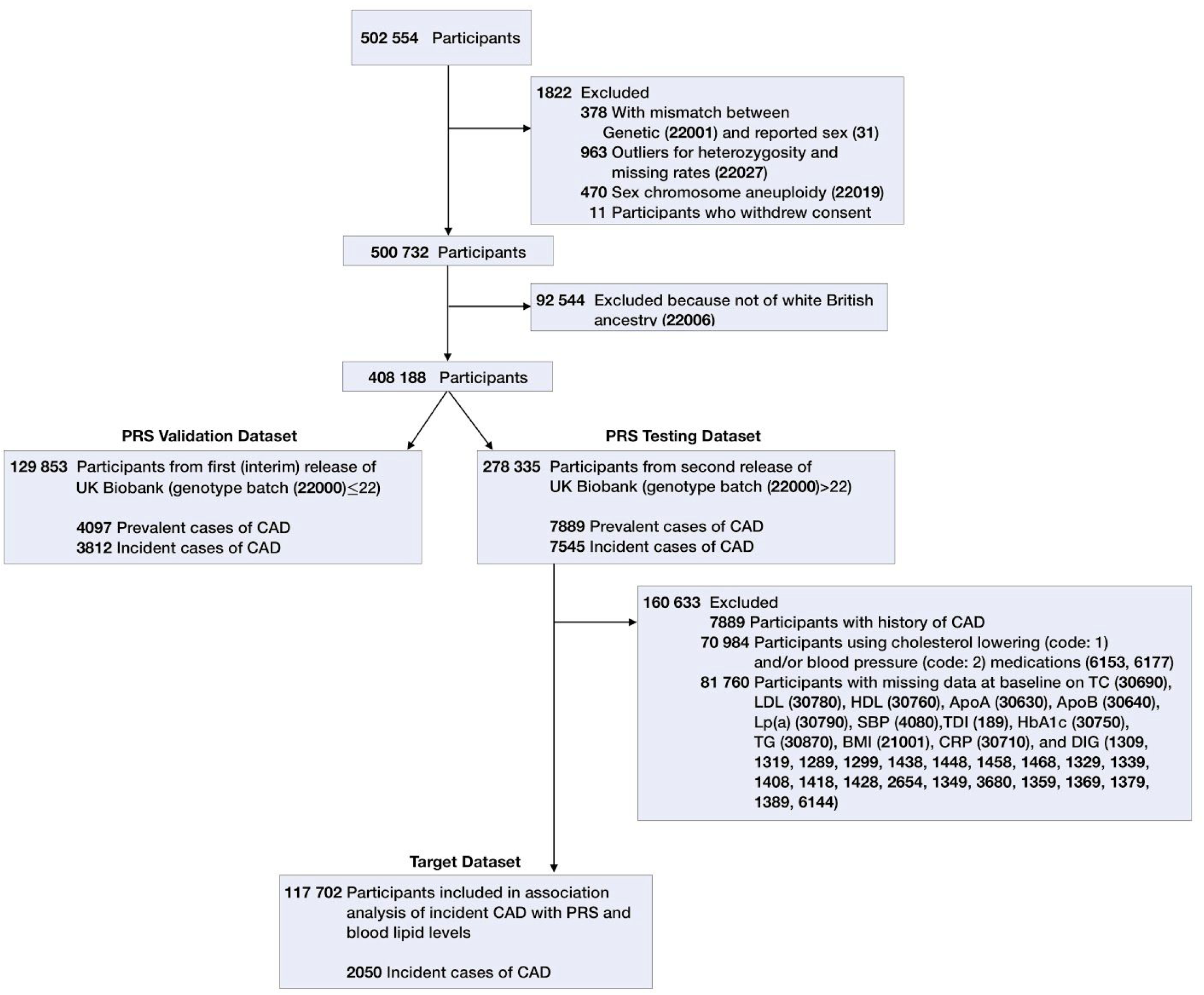
Study design. Flowchart describing the different sub-populations from UK Biobank used in the different steps of the analyses. Numbers in brackets represent UK Biobank data fields used for filtering.

### Development of CAD PRS

To build a new CAD PRS, we applied the stacked clumping and thresholding (SCT) algorithm developed by Privè et al. (2019) to GWAS summary statistics of CAD (Nikpay et al., 2015) together with a validation dataset. This led to the identification of a set of SNPs and effect sizes that best predicts the onset of CAD. We used the interim release of the UK Biobank as a validation dataset, selecting a total of 129,853 individuals with European ancestry (**Figure 1**).

To compare the predictive performance of this new SCT PRS to previously published PRSs, we computed the PRSs from Khera et al. (2018) and Inouye et al. (2018), and built an additional two novel PRSs by combining the PRS algorithm from Khera et al. with SCT (SCT-K) and Inouye et al. with SCT (SCT-I). We assessed the predictive performance of the five PRSs by logistic regression and the Area Under the Curve (AUC) of the Receiver Operating Characteristic (ROC) in a testing dataset comprising 278,335 individuals of European ancestry from the second release of UK Biobank (**Figure 1**). Logistic regression models used PRSs as predictive variables and presence/absence of a CAD outcome and were adjusted for age at recruitment, gender, genotyping array, and the first four principal components of ancestry.

We defined CAD outcome based on self-reported diagnoses and Hospital Episode Statistics (HES) data (see **eTable 1**). Prevalent and incident CAD cases were identified by extracting for each individual of the validation and testing datasets the date of the first CAD event. The prevalent and incident conditions were attributed to CAD events that occurred before or after the date of the first UK Biobank assessment, respectively. In order to adhere with and compare to previously published AUC results (Khera et al., 2018), we used only prevalent CAD cases of the testing dataset for AUC calculation.

**Table 1:** Predictive performance of PRSs for CAD assessed in this study.

| PRS panel | SNPs in PRS | AUC (95% CI) |
| --- | --- | --- |
| Khera | 6630150 | 0.811 (0.802-0.820) |
| Inouye | 1745180 | 0.812 (0.803-0.822) |
| SCT | 291969 | 0.814 (0.806-0.822) |
| SCT+Khera<br>(SCT-K)* | 6699370 | 0.814 (0.806-0.823) |
| SCT+Inouye<br>(SCT-I)* | 1920136 | 0.817 (0.809-0.826) |

Khera: PRS developed by Khera et al. (2018). Inouye: PRS developed by Inouye et al. (2018). SCT: PRS for CAD developed with the SCT algorithm. SCT+Khera (SCT-K): combination of SCT and Khera PRSs. SCT+Inouye (SCT-I): combination of SCT and Inouye PRSs. \* For SNPs present in both SCT and a second panel (khera or Inouye), effect sizes from SCT were taken. AUC (95% CI): Area Under the ROC Curve and 95% confidence intervals.

### Statistical analysis

Statistical analyses have been performed on a subset of the testing dataset comprising incident CAD cases only (**Figure 1**). Exclusion criteria comprised previous history of CAD (7889), use of cholesterol lowering and/or blood pressure medications (70,984), and missing values at baseline on at least one of the following risk factors (81,760): total (TC), high-density (HDL), and low-density (LDL-C) lipoprotein cholesterol, Apolipoprotein A (ApoA), Apolipoprotein B (ApoB), Lipoprotein a (Lp(a)), systolic blood pressure (SBP), Glycated haemoglobin (HbA1c), Triglycerides (TG), Body Mass Index (BMI), C-reactive protein (CRP), dietary intake goals (DIG) and townsend deprivation index (TDI). DIG refers to a list of 10 ideal intake goals (Mozaffarian, 2016) comprising the following diet components: Fruit, Vegetables, Whole grains, (Shell)fish, Dairy products, Vegetable oils, Refined grains, Processed meats, Unprocessed meats, and Sugar-sweetened beverages. This led to a target dataset for statistical analysis of 117,702 individuals, comprising 2050 incident CAD cases, with a median follow-up of 8.2 years (5^th^ - 95^th^ percentiles: 6.2 - 9.4) (**Figure 1**).

Logistic regression was used to determine the ability of PRS and of each lipid risk factor to stratify the risk of incident CAD among individuals of the target dataset. Specifically, the distribution of each risk factor and of PRS was divided into percentiles. Average values of percentiles were used as the predictive variable in logistic regression models to calculate the predicted probability of incident CAD. Logistic regression models were conditioned on the mean value of additional covariates, namely age at recruitment, gender, genotyping array, the first four principal components of ancestry, SBP, family history of heart disease, smoking and diabetes status, TDI, BMI, HbA1C, TG, CRP, and DIG. Odds ratio per percentile of risk factor and PRS distributions were calculated by dividing predicted percentile-dependent CAD probabilities by the probability of the 50^th^ percentile.

In order to assess the correlation of quantitative CAD risk factors (LDL-C, HDL, TC, TC:HDL, LDL-C:HDL, ApoA, ApoB, ApoA:ApoB, Lp(a), BMI, SBP, HbA1C, TG, CRP) and PRS among individuals of the target dataset, a pairwise correlation matrix was calculated. Specifically for each individual, his corresponding percentile value was calculated in each risk factor distribution. Then, a matrix of Pearson correlation coefficients was calculated for each pair of risk factors.

Multivariable Cox regression analyses were performed to assess the association of polygenic risk and different blood lipid levels with incident events of CAD. Blood lipids considered were TC, HDL, LDL-C, as well as TC:HDL and LDL-C:HDL ratios. We divided individuals into three groups based on their polygenic risk: Low PRS, Intermediate PRS, High PRS: <=10th, >10th <=90th, >90th percentile of PRS distribution, respectively. We binned Low PRS, Intermediate PRS, and High PRS populations according to different ranges for each blood lipid and calculated for each bin the Hazard ratio (HR) relative to the Intermediate PRS with Optimal blood lipid levels, used as reference populations. Optimal blood lipid levels were as follows: TC: below 200 mg/dL; LDL-C: below 130 mg/dL; HDL: above or equal to 80 mg/dL; TC:HDL: below 3.0; LDL-C:HDL: below 2.0.

In order to assess for possible interactions between LDL-C and PRS and between HDL and PRS, we performed a Cox regression analysis with quantitative scaled PRS, scaled LDL-C, scaled HDL, LDL-C×PRS and HDL×PRS as variables.

All Cox regression analyses were adjusted for age at recruitment, gender, genotyping array, the first four principal components of ancestry, SBP, family history of heart disease, smoking and diabetes status, TDI, BMI, HbA1C, TG, CRP, and DIG. The DIG variable has been modelled as an integer variable ranging from 0 to 10, reflecting the number of dietary intake goals achieved by each participant. UK Biobank fields used to model the DIG variables were taken from Said et al. (2018). To maximize the likelihood of reporting true findings, we set a P value threshold for significance at 0.005 instead of 0.05 (Benjamin et al., 2018).

## Results

### Predictive performances of CAD PRSs

The SCT algorithm identified a set of 291,969 SNPs that best predicted the onset of CAD in the validation dataset (**Figure 1**). Since SCT identified a smaller number of SNPs than PRSs from Khera (Khera et al., 2018; 6,630,150 SNPs) and Inouye (Inouye et al., 2018; 1,745,180 SNPs), we assessed if the additional SNPs in these two PRSs may have residual predictive power in combination with STC-selected SNPs. We therefore generated and tested two additional PRSs by combining PRS from Khera et al. with SCT (SCT-K; 6,699,370 SNPs) and Inouye et al. with SCT (SCT-I; 1,920,136 SNPs).

**Table 1** shows that the new SCT PRS displayed higher predictive performance than PRSs from Khera et al. and Inouye et al. Moreover, while the addition of the SNPs from Khera PRS to SCT (i.e., SCT-K PRS) didn’t result in any significant improvement in predictive performances over SCT alone, the combination of SCT and Inouye SNPs (SCT-I PRS) generated a new PRS with the highest predictive performances. We thus used the SCT-I PRS in the following analyses.

### Odds Ratio of PRS and blood lipids

We next examined the ability of SCT-I PRS to stratify the risk of CAD in the UK Biobank population and compared that with those of blood lipids and associated (Apo)lipoproteins recorded at baseline. In order to avoid reverse causation, we considered only incident cases of CAD. The target population used for the analysis (Baseline characteristics are provided in **Table 2**) was generated from the testing dataset, by excluding prevalent CAD cases, as well as all individuals with missing data on TC, HDL, LDL-C, ApoA, ApoB, Lp(a), SBP, HbA1c, TG, BMI, CRP, and DIG at baseline (**Figure 1**). **Figure 2** shows the odds ratio per percentile of blood lipid and PRS distributions, computed respect to the average of each distribution.

**Table 2:** Baseline characteristics of the target population.

| Characteristics | Males | Females | Low PRS | Intermediate PRS | High PRS |
| --- | --- | --- | --- | --- | --- |
| Total, N° (%) | 49476 (42.03) | 68226 (57.9) | 11770 (10) | 94161 (80) | 11771 (10) |
| Age, mean (SD), y | 55.15 (8.19) | 55.43 (7.98) | 55.93 (7.98) | 55.33 (8.08) | 54.52 (8.07) |
| CAD cases, N° (%) | 1462 (2.96) | 588 (0.86) | 93 (0.79) | 1542 (1.64) | 415 (3.53) |
| LDL-C, mean (SD), mg/dL | 144.61 (29.55) | 143.66 (31.85) | 138.31 (30.16) | 144.17 (30.81) | 148.93 (31.55) |
| TC, mean (SD), mg/dL | 224.48 (38.28) | 231.40 (40.99) | 222.37 (39.29) | 222.36 (39.9) | 233.54 (40.81) |
| HDL, mean (SD), mg/dL | 50.81 (11.63) | 62.66 (13.83) | 58.82 (14.49) | 57.66 (14.19) | 56.69 (14.01) |
| LDL-C:HDL, mean (SD) | 2.98 (0.85) | 2.41 (0.77) | 2.50 (0.81) | 2.65 (0.85) | 2.78 (0.87) |
| TC:HDL, mean (SD) | 4.60 (1.12) | 3.84 (0.98) | 3.97 (1.05) | 4.16 (1.11) | 4.32 (1.13) |
| ApoA, mean (SD), g/L | 1.44 (0.22) | 1.65 (0.26) | 1.58 (0.27) | 1.56 (0.27) | 1.54 (0.27) |
| ApoB, mean (SD), g/L | 1.08 (0.22) | 1.05 (0.23) | 1.02 (0.22) | 1.06 (0.23) | 1.10 (0.23) |
| ApoB:ApoA, mean (SD) | 0.77 (0.20) | 0.66 (0.19) | 0.66 (0.19) | 0.70 (0.20) | 0.74 (0.21) |
| Lp (a), mean (SD), nmol/L | 42.54 (48.39) | 44.28 (49.16) | 33.35 (40.12) | 42.86 (48.24) | 59.23 (57.24) |
| Smoking, N° (%) | 5598 (11.31) | 5015 (7.35) | 984 (8.36) | 984 (9.00) | 11.59 (9.84) |
| Family history, N° (%) | 18205 (36.80) | 29490 (43.22) | 4125 (35.05) | 37978 (40.33) | 5592 (47.51) |
| SBP , mean (SD) mmHg | 139.44 (16.98) | 133.13 (18.68) | 133.98 (17.77) | 135.83 (18.26) | 137.19 (18.56) |
| Diabetes, N° (%) | 266 (0.54) | 206 (0.30) | 36 (0.31) | 385 (0.41) | 51 (0.43) |
| BMI, mean (SD), kg/m<sup>2</sup> | 27.04 (3.80) | 26.28 (4.66) | 26.38 (4.21) | 26.60 (4.33) | 26.78 (4.49) |
| HbA1C, mean (SD), mmol/mol | 34.76 (4.99) | 34.71 (4.21) | 34.68 (4.36) | 34.73 (4.53) | 34.79 (4.89) |
| TG, mean (SD), mg/dL | 169.92 (95.56) | 130.10 (69.35) | 141.11 (79.30) | 147.01 (83.81) | 151.19 (87.11) |
| CRP, mean (SD), mg/L | 2.25 (4.10) | 2.40 (4.04) | 2.28 (4.15) | 2.34 (4.02) | 2.41 (4.32) |

**Figure 2.**
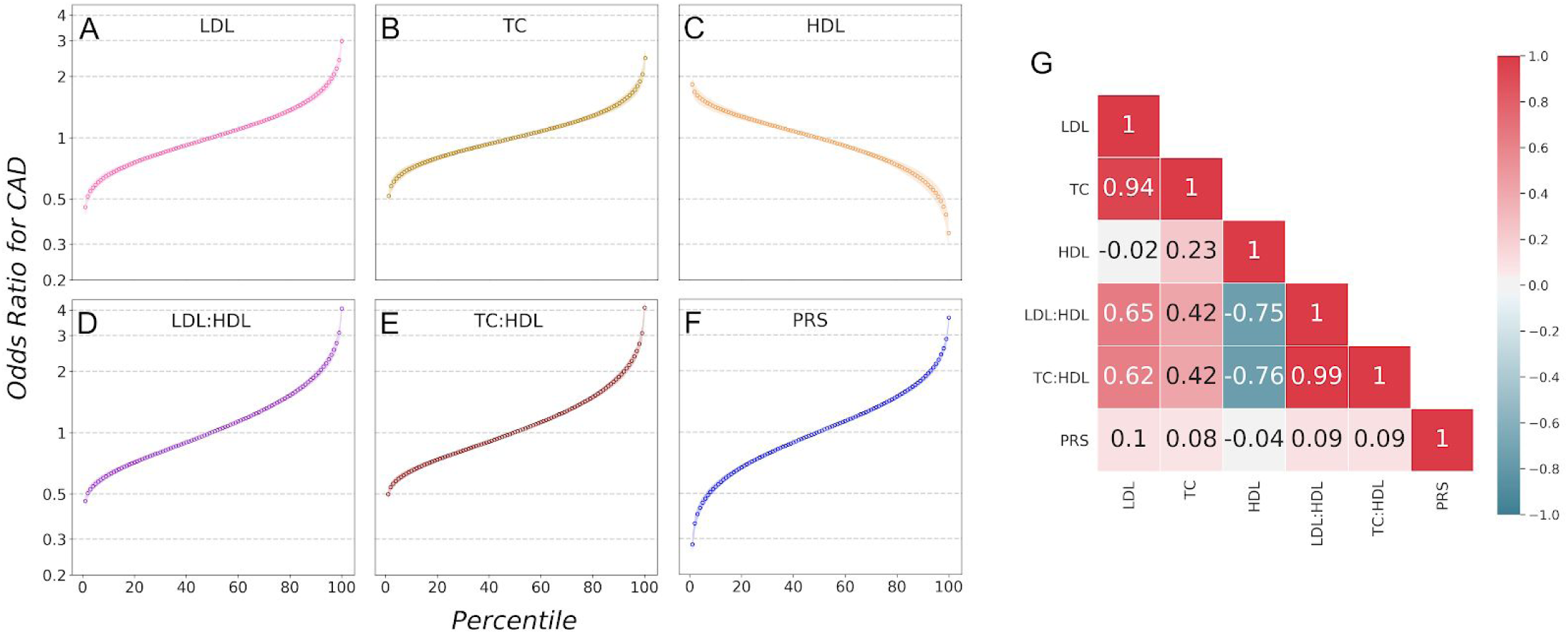
Correlation matrix and odds ratio for incident CAD by PRS, lipids and their ratios. Predicted odds ratio of incident CAD for each percentile of the distribution of the following CAD risk factors: LDL-C (**Panel A**), TC (**Panel B**), HDL (**Panel C**), LDL-C:HDL ratio (**Panel D**), TC:HDL ratio (**Panel E**), and SCT-I PRS (**Panel F**). Odds ratio were calculated through logistic regression models with percentiles-specific average values of PRS and risk factors as predictive variables and incident CAD outcomes as response variables. **Panel G**. Matrix of pearson correlation coefficients highlighting the pairwise correlation between distributions of blood lipid concentrations and PRS score in the target dataset population.

Lipid ratios (TC:HDL and LDL-C:HDL) as well as SCT-I PRS displayed wider odds ratio ranges than single blood lipids alone (TC, LDL-C, and HDL): odds ratio for TC:HDL, LDL-C:HDL, and PRS ranged respectively from 0.50 to 4.12, from 0.46 to 4.07, and from 0.28 to 3.56, across percentiles of the corresponding distributions. On the other hand, odds ratio for TC, LDL-C, and HDL displayed narrower ranges, from 0.52 to 2.46, from 0.46 to 2.99, and from 1.83 to 0.34, respectively.

Similar results were obtained for (Apo)lipoproteins (see **eFigure 1**), where the odds ratio range of ApoB:ApoA (from 0.46 to 4.15) was wider than that of ApoB (from 0.42 to 3.35) and ApoA (from 1.76 to 0.41) alone. Finally, Lp(a) odds ratio range was the least observed (from 0.95 to 1.83).

Notably, among the factors with wider odds ratio range, TC:HDL, LDL-C:HDL, ApoB:ApoA, and PRS, the latter generated the wider range, with a 13 fold increase in odds ratio between the bottom and the top percentiles of the distribution, compared to a 8.25, 8.82, and 9.05 fold increase for TC:HDL, LDL-C:HDL, and ApoB:ApoA, respectively.

### Correlation between PRS and traditional risk factors

We then investigated if PRS and traditional risk factor distributions are correlated among individuals of the target population. **Figure 1G** displays the correlation matrix. Correlation coefficients displayed high (from ±0.50 to ±1.00) to moderate (from ±0.30 to ±0.49) degrees of correlation when any pair of blood lipids distributions were involved, the only exception being the low correlation between LDL-C and HDL. Most notably, the correlation coefficients between PRS and any blood lipids were very low, ranging from −0.04 for PRS with HDL to 0.1 for PRS to LDL-C. This finding indicates that there is a very low degree of overlap between individuals stratified through PRS and through any blood lipids, suggesting that PRS and blood lipids capture different components of the CAD risk.

Similar low values of correlation for PRS were obtained when additional risk factors are compared, such as ApoA, ApoB, ApoB:ApoA ratio, Lp(a), BMI, and SBP, HbA1C, TG, and CRP (see **eFigure 2**). CRP was moderately correlated only with BMI (0.44), possibly being related to inflammation processes in overweight individuals (Visser et al. 1999). TG were moderately correlated with each lipid, apolipoprotein and BMI, and highly correlated with their ratios (TC:HDL = 0.69, LDL:HDL = 0.64, ApoB:ApoA = 0.52).

### Association of lipid levels and polygenic risk with incident CAD

PRS and blood lipids were orthogonal, capturing different independent components of CAD risk. It was of interest to investigate how the interplay between these two orthogonal sources contribute to the overall CAD risk in the population. **Figure 3** illustrates the results of a Cox regression analysis testing the association of different lipid levels with incident CAD in three populations: individuals with a high polygenic risk (High PRS: individuals at the top 10 percentiles of PRS distribution), individuals with intermediate polygenic risk (Intermediate PRS: between the bottom 10 and the top 10 percentiles of PRS distribution), and individual with low polygenic risk (Low PRS: below the bottom 10 percentiles of the PRS distribution).

**Figure 3.**
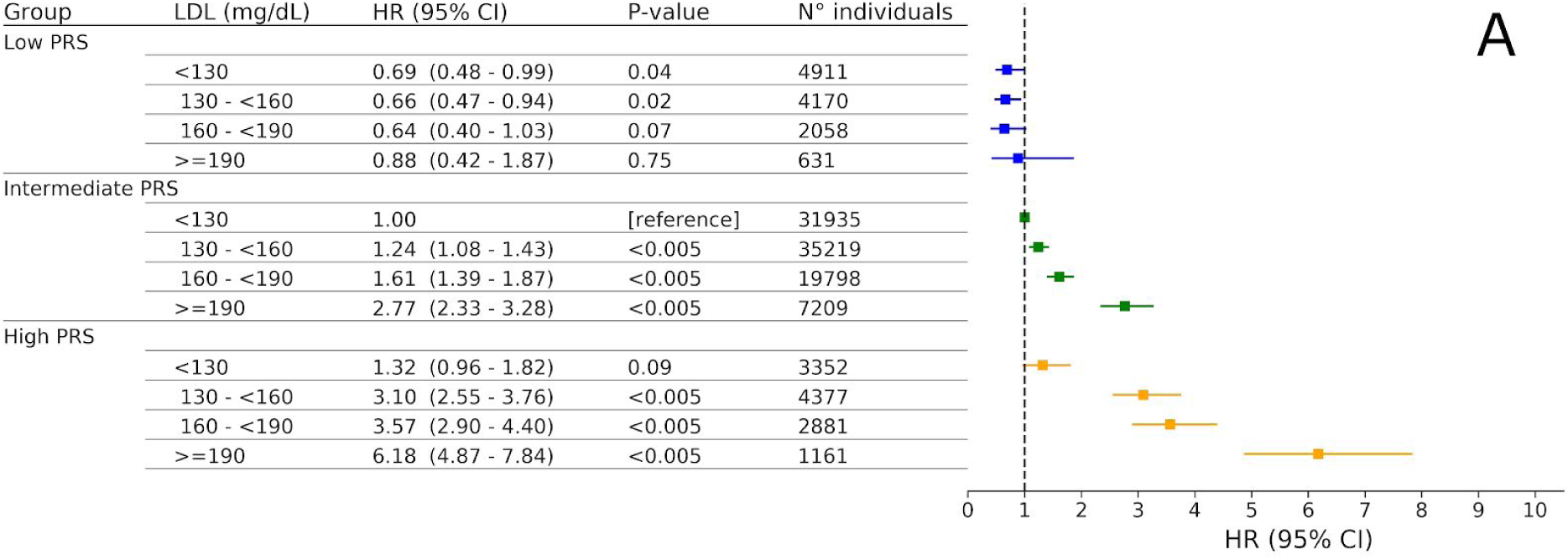
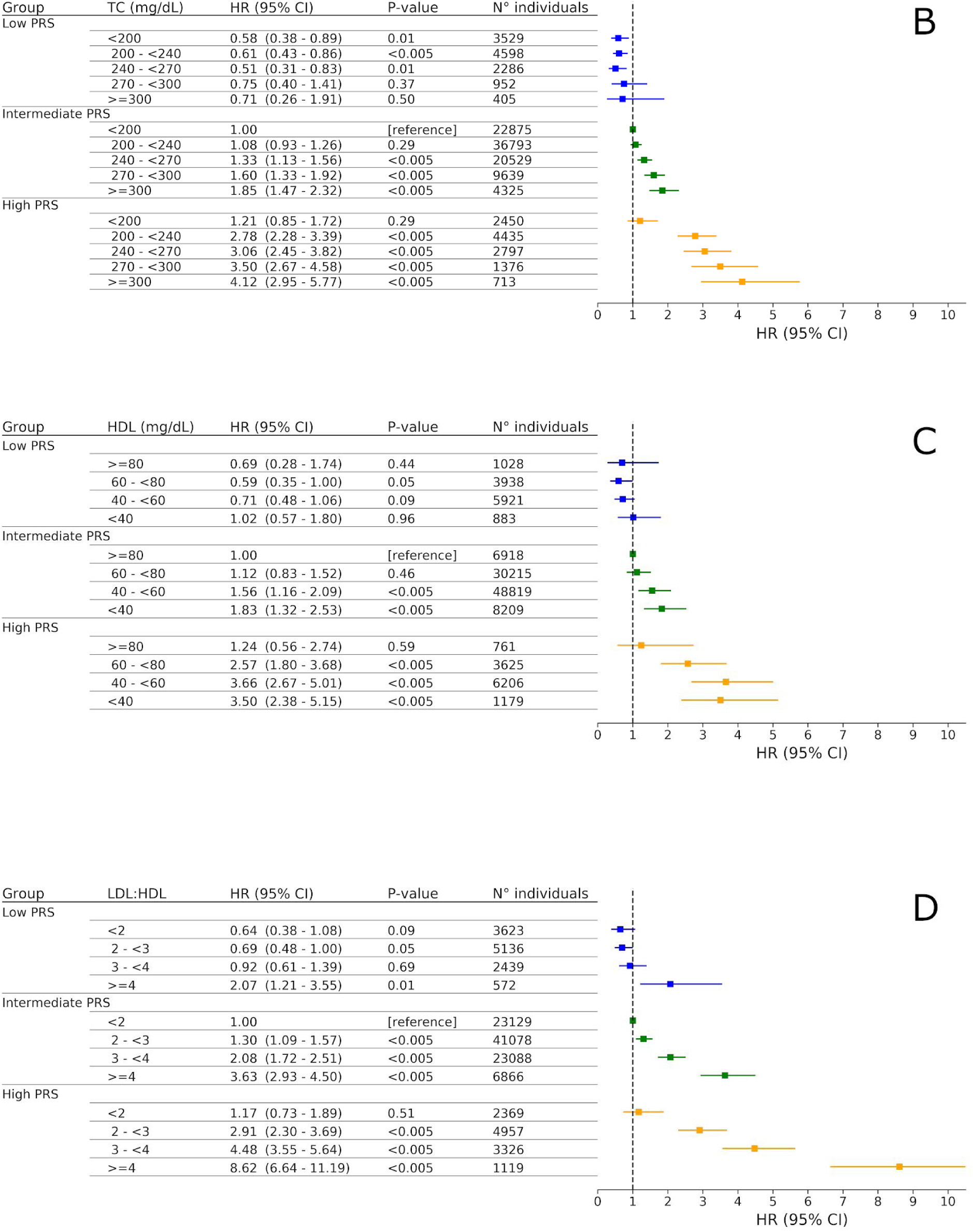
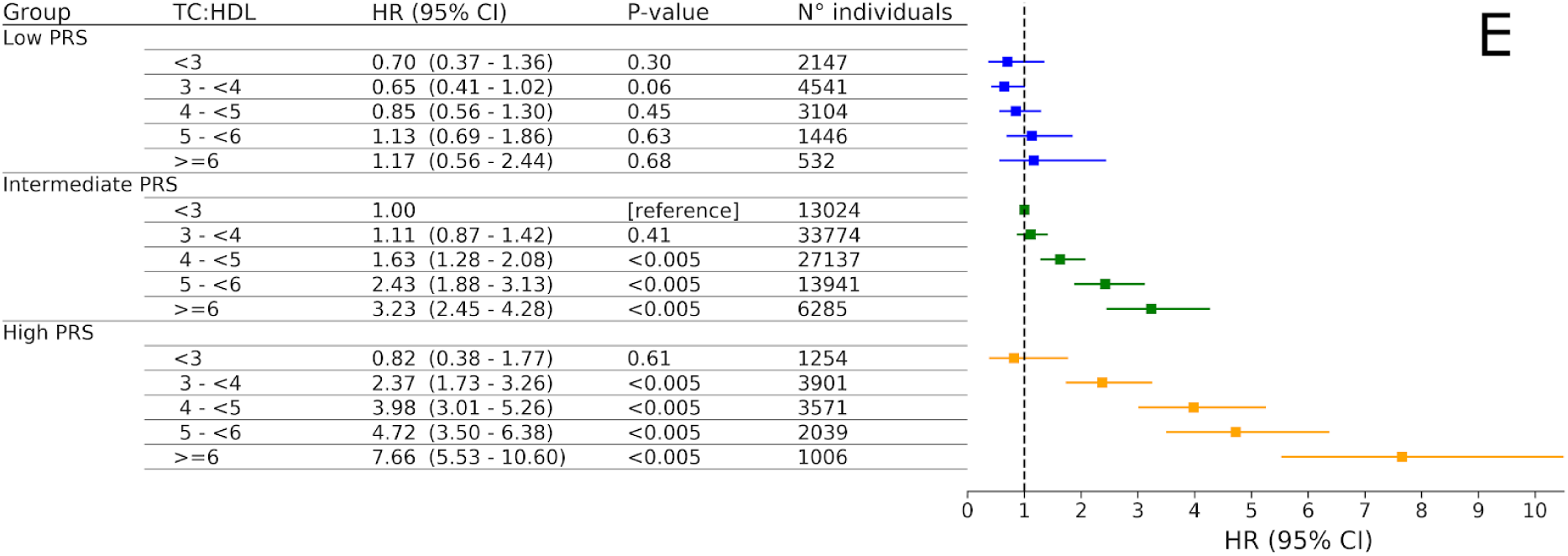
Risk of CAD events in the UK Biobank prospective cohort according to polygenic risk and blood lipid levels. The population of the target dataset was divided in three groups: Individuals with Low (bottom 10 percentiles of PRS distribution), Intermediate (between the bottom 10 and the top 10 percentiles of PRS distribution), and High (above the top 10 percentiles of PRS distribution) PRS. These three groups were stratified according to different ranges of LDL-C (**Panel A**), TC (**Panel B**), HDL (**Panel C**), LDL-C:HDL (**Panel D**), and TC:HDL (**Panel E**) levels. For each blood lipid, HR were calculated by comparing each lipid level of Low, Intermediate, and HIgh PRS groups relative to the reference population in a Cox proportional-Hazards model adjusted for age, gender, genotyping array, the first 4 principal components of ancestry, townsend deprivation index, diabetes and smoking status, family history of heart disease, systolic blood pressure, Glycated haemoglobin, Triglycerides, Body Mass Index, C-reactive protein, and dietary intake goals. For each lipid, the reference population corresponded to the first lipid level in the Intermediate PRS subpopulation. For each lipid level, the corresponding population size is reported (N° Individuals). The vertical dashed line indicates the reference HR value of 1.

For LDL-C (**Figure 3A**) above 130 mg/dL, HRs increase for higher levels, with HRs of High PRS being at least two fold higher than those of Intermediate PRS. Below the 130 mg/dL threshold, HR in High PRS group was not statistically significantly higher compared to Intermediate PRS with the same LDL-C level. High PRS with borderline LDL-C (130-159 mg/dl) group showed higher HR (3.10; 95% CI, 2.55-3.76) than Intermediate PRS with very high LDL-C (>190 mg/dl; HR 2.77; 95% CI, 2.33-3.28). On the other hand, in the Low PRS group the highest LDL-C level (> 190 mg/dl) did not show statistically significant increased risk compared to optimal LDL-C level (<130 mg/dl) in Intermediate PRS. Notably, relative risk reduction between Very High and Optimal LDL-C levels were lower in Low PRS (45%) compared to High PRS (618%) and Intermediate PRS (277%).

Similar behavior was observed for TC (**Figure 3B**): In High PRS, TC levels displayed a positive association with CAD for values above 200 mg/dL, while there was not significant association at TC <200 mg/dL. Incident CAD risk was inversely associated with HDL levels (**Figure 3C**). In High PRS, a positive association with incident CAD was observed for HDL levels below 80 mg/dL. In High and Intermediate PRS, Lipid ratios (LDL-C:HDL and TC:HDL in **Figure 3D** and **3E**, respectively) showed a positive association with CAD for values above 2.0 and 3.0 for LDL-C:HDL and TC:HDL, respectively. In High PRS, lipid ratios displayed a much stronger positive association with incident CAD compared to single lipid levels. Despite this, in High PRS individuals Optimal values of lipid ratios (below 2.0 and 3.0 for LDL-C:HDL and TC:HDL, respectively) displayed not statistically significant HRs compared to Intermediate PRS with the same Optimal values. Very High LDL-C:HDL were positively associated with CAD incidence even in the Low PRS group (HR 2.07; 95% CI, 1.21-3.55).

### Interactions between LDL-C, HDL and PRS with incident CAD

In order to assess for possible interactions between LDL-C and PRS and between HDL and PRS, we performed a Cox regression analyses with quantitative scaled PRS, scaled LDL-C, scaled HDL, LDL-CxPRS and HDLxPRS as variables controlling for lifestyle and clinical factors and biomarkers associated with CAD incidence (see Methods for details). According to a P value threshold for significance of 0.005, we identified significant interactions between LDL-C and PRS (HR: 1.07, P value: <0.005, **eTable 2**), but not between HDL and PRS (HR: 0.94 P value: 0.02, **eTable 2**). We next assessed the HRs of PRS, LDL-C and their interaction (LDL-CxPRS) excluding from the model the non-significant interaction between PRS and HDL (HDLxPRS). PRS showed higher HR per SD (1.45; 95% CI, 1.39-1.53) than LDL-C (HR 1.39; 95% CI, 1.32-1.45) and their interaction (LDL-CxPRS) had a p-value lower than 0.005 (**eTable 3)**.

In order to explore the effect of the above findings, we assessed the association with incident CAD for different levels of LDL-C and HDL in each polygenic risk group separately, taking as reference Optimal LDL-C (<130 mg/dL) and Very High HDL (>80 mg/dL) (**eFigure 3** and **4**), respectively. While for LDL-C, High PRS group showed much steeper increase in HR than Low PRS and Intermediate PRS groups for increasing LDL-C, for HDL the polygenic risk groups showed not significantly increased HRs across HDL levels tested.

## Discussions

Here, we analyzed the interplay between PRS and clinical blood lipids in determining the risk of incident CAD in the prospective cohort of the UK Biobank. The analysis of the association of incident CAD with different lipid levels and our novel PRS in three subpopulations at different polygenic risk (Low: bottom decile, Intermediate: 10th-90th percentiles and High: top decile) supports the following conclusions. First, we have identified for the first time a significant interaction between polygenic risk and LDL-C in determining CAD outcome. The effect that LDL-C levels have on an individual’s CAD risk depends on the polygenic background. Second, we find that the combination of High PRS and Borderline LDL-C gives individuals an incident CAD risk that is greater than individuals with Intermediate or Low PRS but who have Very High LDL-C (>190mg/dL). Third, individuals with Optimal lipid levels do not show increased CAD risk despite their High polygenic risk. Fourth, individuals in the lowest 10 percentiles of the PRS distribution are not at increased risk of CAD regardless of their LDL-C levels.

These results have important implications for the primary prevention of CAD. For example, according to current guidelines (Stone et al., 2014), individuals with Borderline LDL-C (130-159 mg/dL) will not be recommended statins. However according to our results, individuals in this group who also have High polygenic risk have a greater relative risk of CAD than individuals with Intermediate or Low polygenic risk, but whose Very High LDL-C (>190 mg/dL) would lead to the recommendation of statins. Our analysis also shows that the higher risk of incident CAD in High PRS individuals is reduced to that of the reference population if lipid levels are within the following conditions: <130 mg/dL for LDL-C, <200 mg/dL for TC, >80 mg/dl for HDL, and <2.0 and 3.0 for LDL-C:HDL and TC:HDL, respectively.

A prior study (Christiansen et al., 2020) identified a strong positive association between total plaque burden and a CAD PRS, with an increase of +78% of coronary artery calcium scores and a +16% of severe stenosis score per SD increase in PRS. Notably, the levels of LDL-C - that is the blood lipid mostly contributing to plaque formation (Bentzon et al., 2014) - were very similar in all polygenic risk groups, (average LDL-C: 120 mg/dL, 128 mg/dL, and 131 mg/dL in Low, Average, and High PRS, respectively). This is consistent with the finding of our study, suggesting that specific polygenic backgrounds may moduate the atherosceltric effect of LDL-C.

To our knowledge, this is the first report that extensively compared the odds ratio range of a PRS (SCT-I) with a wide set of blood lipids and Apolipoproteins. We observed that each lipid (and apolipoproteins) ratios generated wider odds ratio ranges across the percentiles of their distributions than single lipids (Figure 2, eFigure 1). This agrees with previous work highlighting that TC:HDL and LDL-C:HDL display higher predictive performances for CAD than single lipids (Shai et al., 2004; Ingelsson et al., 2007). In addition, we showed that PRS on its own has a greater odds ratio range than any individual blood lipid and their ratios (Figure 2).

We also reported for the first time the association of lipid levels to incident CAD in groups with different polygenic risk. However similarly to our analyses, two previous studies (Khera et al., 2016; Said et al., 2018) analyzed the interplay between different polygenic backgrounds with a modifiable parameter: lifestyle (poor/unfavorable, intermediate, and ideal/favorable). In both studies, lifestyle was modelled as an aggregated categorical variable comprising smoking status, BMI, frequency of physical activity, and ideal or poor diet. These studies and our results share similar patterns of interplay between the polygenic background and the additional factor considered. That is, they all observe a monotonic increase of CAD association (HRs) with increasing polygenic risks and the additional factor (be it lifestyle or lipid levels). However, there are two notable differences between this study and those mentioned above. First, in our analysis even in individuals at High polygenic risk, Optimal lipid level thresholds show non significant increased risk compared to Intermediate polygenic risk with Optimal blood lipids. This is not observed in either Khera et al. (2016) or Said et al. (2018), where the CAD risk in High PRS individuals is never completely reduced in favourable lifestyles categories. Second, Said et al. (2018) found no significant interaction between lifestyle and genetic risk with CAD. In contrast, we have identified a statistically significant interaction between PRS and LDL-C in modulating individual’s risk of CAD.

A previous study (Hindy et al., 2018), analyzing the association of smoking status to incident CAD in different polygenic backgrounds (lower, intermediate, and upper quartiles of PRS distribution) identified statistically significant interaction between smoking and PRS, highlighting the importance of considering the polygenic background in studying lifestyle effects on CAD risk.

Elliott and collegues (2020) showed that a model using age, sex and PRS as covariates to predict CAD events had the same predictive power (C-statistics 0.76) of the Pooled Cohort Equation (PCE), a widely used model comprising several risk factors such as TC, HDL, age, sex, SBP, smoking, diabetes status and ethnicity (Yadlowsky et al. 2016). PRS has the advantage to be detected early in life, as opposed to conventional risk factors used in the PCE that become relevant at mid and late age. Therefore PRS can allow the implementation of preventive strategies earlier in life. The potential efficacy of early intervention in the primary prevention setting is further supported by the recent evidence (Ference et al. 2019) that a lifelong genetic exposure to 38.67-mg/dL lower LDL-C and 10-mm Hg lower SBP was associated with a 4.5 fold risk reduction of major coronary events. The finding of our study that increased CAD risk conferred by LDL-C is enhanced by the polygenic risk, indicates that individuals with High polygenic risk could benefit the most from lifetime exposure to lower LDL-C levels, warranting the consideration of early detection of both PRS and LDL-C in the general population.

### Strengths and Limitations

Major strengths of this study were: the development and application of a new CAD PRS with higher predictive performance than previously published CAD PRS, a large prospective cohort of about 120 000 individuals, as well as the adjustment for lifestyle (smoking, diet intake goal, BMI), clinical (diabetes, blood pressure, family history), biomarkers (Glycated haemoglobin, Triglycerides, C-reactive protein) and various demographic (TDI) confounders.

This study has limitations. First, lipid levels used in the analyses are those reported at baseline. Therefore, changes in lipid levels over time were not considered in the present analyses. Second, the analysis has been performed on a population of white british individuals. This has implications on the generalizability of the reported results on different ethnic groups or cohorts. Third, the association of different lipid levels with CAD risk cannot be taken as a causal relationship, since lipid levels status were not randomised. Therefore, future research is required to confirm reported results on additional cohort populations and to address the causal relationship between achieving lower lipid levels and reduced CAD risk in the High polygenic risk group.

### Conclusions

In conclusion, physiological parameters, such as blood lipid levels, together with polygenic background, represent two independent contributions to the overall risk of incident CAD. Risk of incident CAD conferred by high LDL-C level was modified by the polygenic risk. However for various lipids, lipid levels at Optimal thresholds showed completely counterbalanced CAD risk despite a High polygenic background. The tailoring of lipid guidelines on the basis of the polygenic background seems a promising strategy to reduce the incidence of CAD in the general population.

## Data Availability

UK Biobank data application n:40692

## Supplemental Tables

**eTable 1:** List of UK Biobank fields and codes defining CAD outcomes.

| **Coronary Artery Disease (CAD)** |  |  |
| --- | --- | --- |
| **UK Biobank Field** | **Description** | **Codes** |
| 20004 | Self Reported Operation code | 1070, 1095,<br>1523 |
| 20002 | Self Self-reported non-cancer illness | 1075 |
| 6150 | Vascular/heart problems diagnosed<br>by doctor | 1 |
| 41270 | Diagnoses - ICD10 | I21X, I22X, I23X,<br>I24X, I252 |
| 40001<br>40002 | Underlying (primary) cause of death:<br>ICD10<br>Contributory (secondary) causes of death:<br>ICD10 | I21X, I22X, I23X,<br>I24X, I252 |
| 41272 | Operative procedures - OPCS4 | K401-K404,<br>K411-K414,<br>K451-K455,<br>K491-K492<br>K498-K499,<br>K502,<br>K751-K754,<br>K758-K759 |

**eTable 2:** Interactions between LDL-C and PRS and between HDL and PRS.

|  | HR(95% CI) | P value |
| --- | --- | --- |
| PRS | 1.40 (1.33 - 1.48) | <0.005 |
| LDL-C | 1.44 (1.37 - 1.51) | <0.005 |
| HDL | 0.75 (0.70 - 0.80) | <0.005 |
| PRS x LDL-C | 1.07 (1.03 - 1.12) | <0.005 |
| PRS x HDL | 0.94 (0.90 - 0.99) | 0.02 |

Interactions between continuous scaled PRS and continuous scaled LDL-C and between PRS and continuous scaled HDL. HRs (95% CIs) refer to a 1 SD variable increase. HRs have been obtained in a Cox proportional-Hazards model adjusted for age, gender, genotyping array, the first 4 principal components of ancestry, townsend deprivation index, diabetes and smoking status, family history of heart disease, systolic blood pressure, Glycated haemoglobin, Triglycerides, Body Mass Index, C-reactive protein, and dietary intake goals.

**eTable 3:**
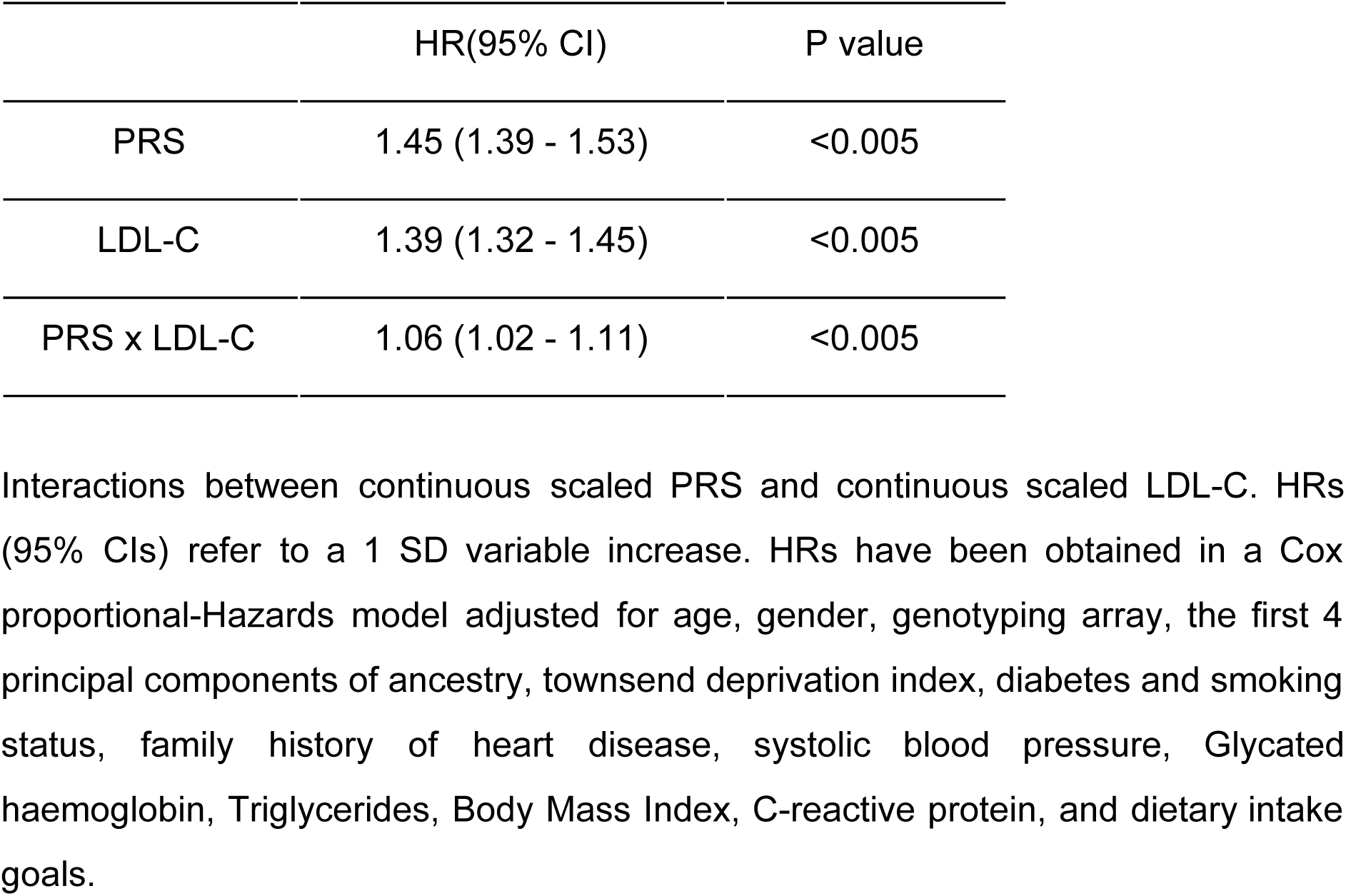
Interactions between LDL-C and PRS.

|  | HR(95% CI) | P value |
| --- | --- | --- |
| PRS | 1.45 (1.39 - 1.53) | <0.005 |
| LDL-C | 1.39 (1.32 - 1.45) | <0.005 |
| PRS x LDL-C | 1.06 (1.02 - 1.11) | <0.005 |

Interactions between continuous scaled PRS and continuous scaled LDL-C. HRs (95% CIs) refer to a 1 SD variable increase. HRs have been obtained in a Cox proportional-Hazards model adjusted for age, gender, genotyping array, the first 4 principal components of ancestry, townsend deprivation index, diabetes and smoking status, family history of heart disease, systolic blood pressure, Glycated haemoglobin, Triglycerides, Body Mass Index, C-reactive protein, and dietary intake goals.

## Supplemental Figures

**eFigure 1.**
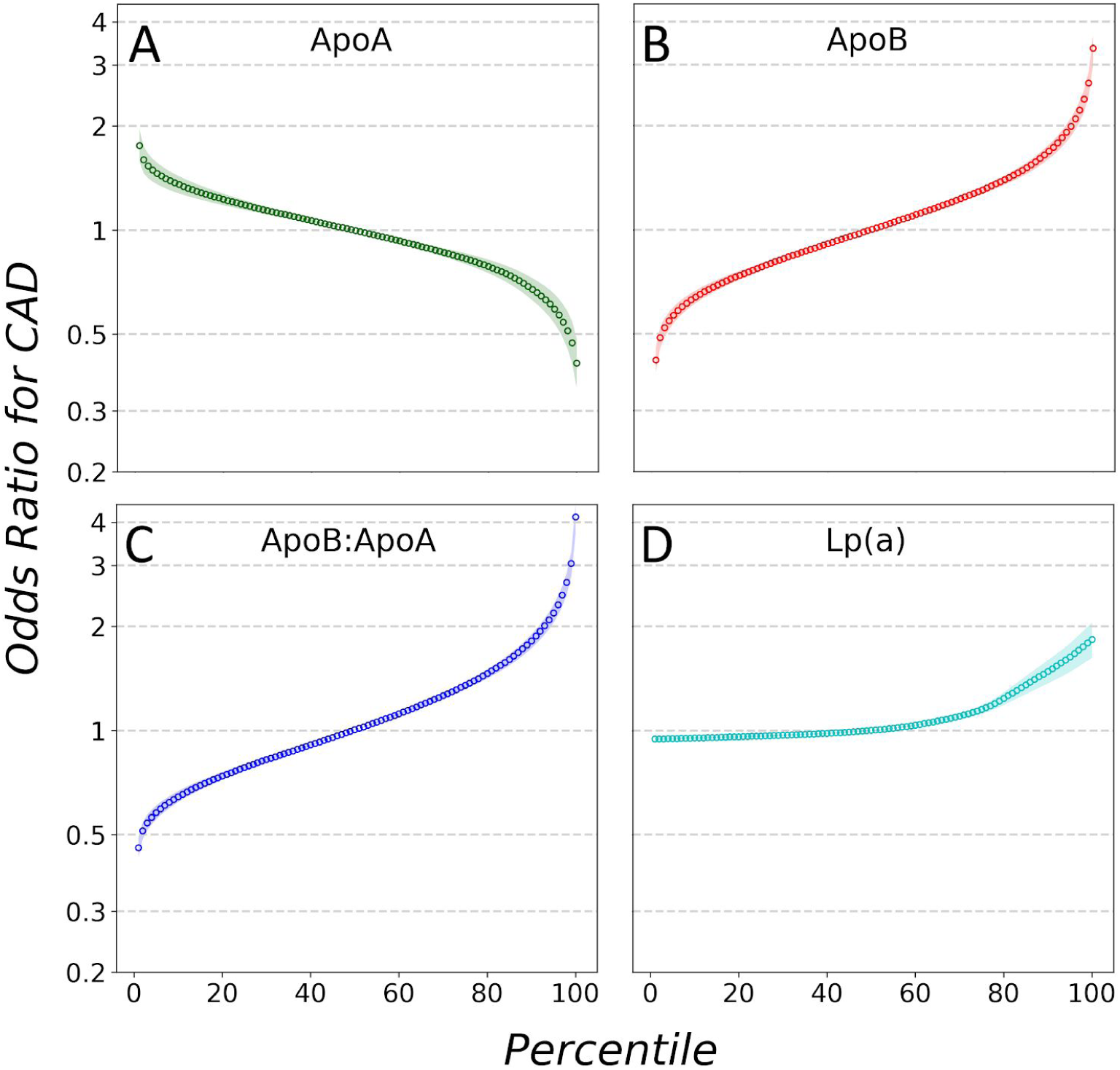
Odds ratio for incident CAD by apolipoproteins. Predicted odds ratio of incident CAD for each percentile of the distribution of the following CAD risk factors: ApoA (**Panel A**), ApoB (**Panel B**), ApoB:ApoA ratio (**Panel C**), and Lp(a) (**Panel D**). Odds ratio were calculated through logistic regression models with percentiles-specific average values of risk factors as predictive variables and incident CAD outcomes as response variables.

**eFigure 2.**
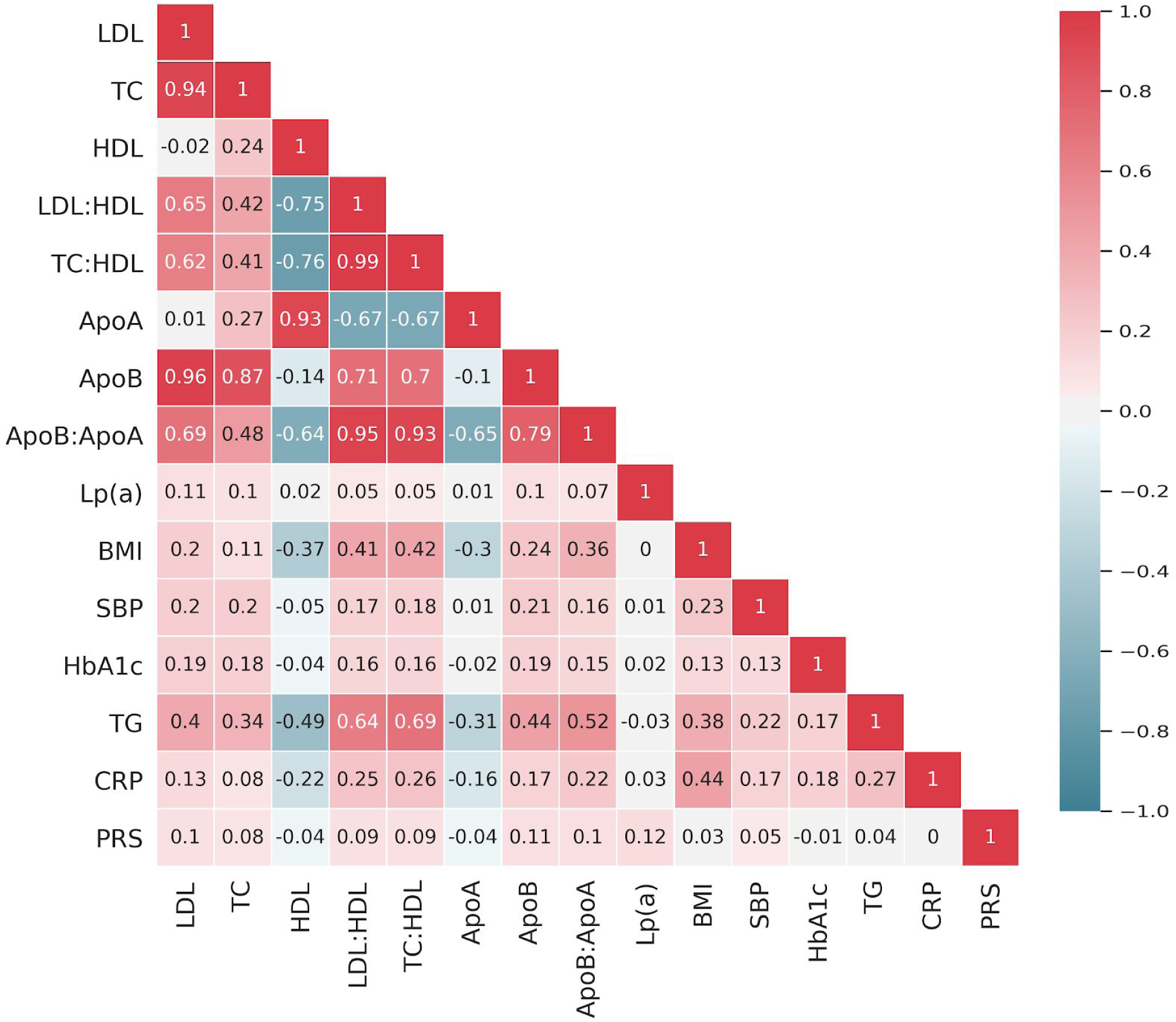
Correlation matrix between CAD risk factors. Matrix of pearson correlation coefficients highlighting the pairwise correlation between distributions of blood lipid concentrations and PRS score in the target dataset. TC: total cholesterol, HDL high-density cholesterol, LDL-C: low-density cholesterol, LDL-C:HDL: low-density over high-density cholesterol ratio, TC:HDL: low-density over high-density cholesterol ratio, ApoA: Apolipoprotein A, ApoB: Apolipoprotein B, Ln(a): Lipoprotein a, BMI: Body Mass Index, SBP: Systolic Blood Pressure, HbA1C: Glycated haemoglobin, TG:Triglycerides, CRP: C-reactive protein, PRS: SCT-I Polygenic risk score.

**eFigure 3.**
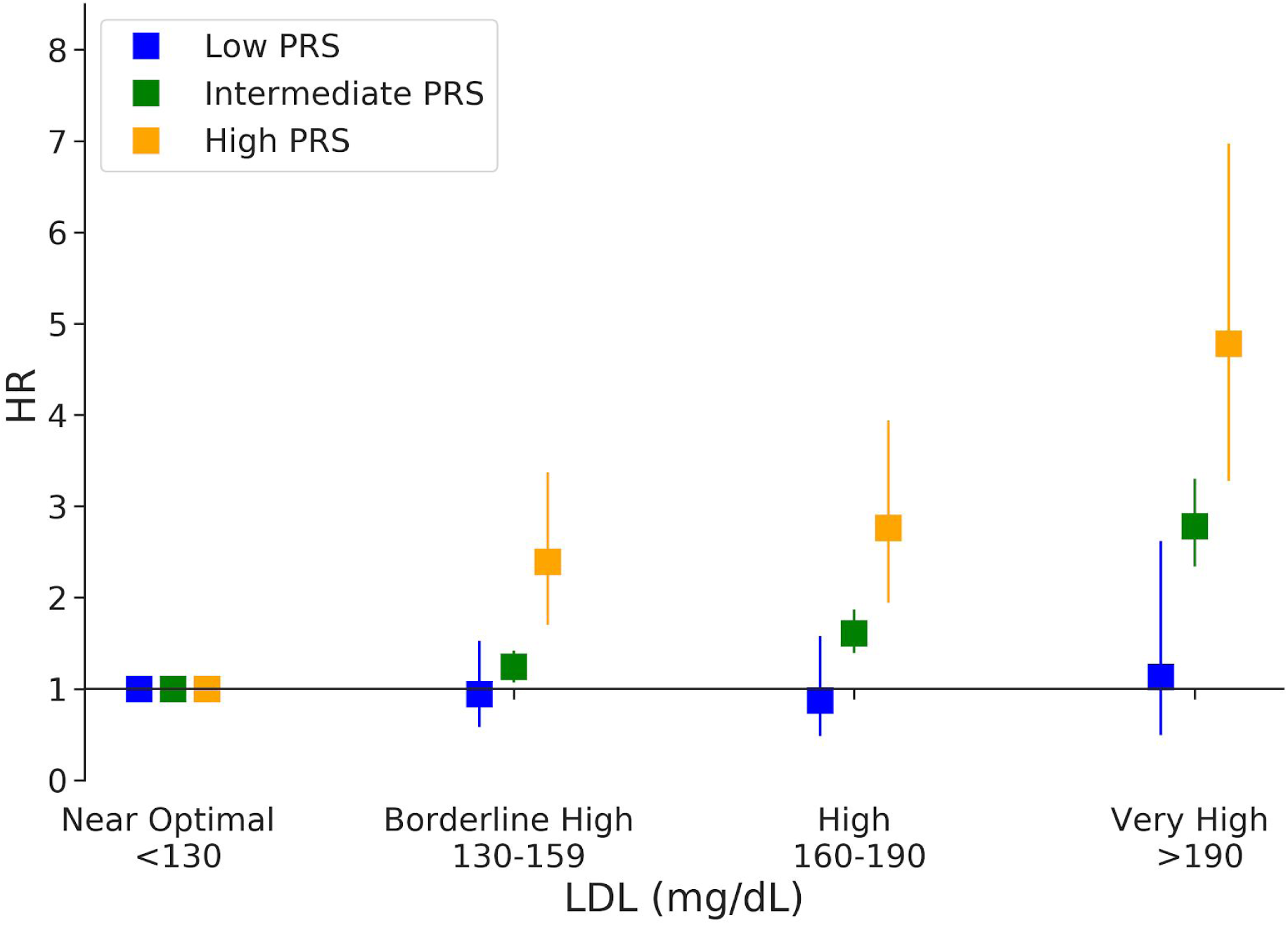
Differential increased CAD risk by LDL-C in groups with different polygenic risk. Increased risk Association of LDL-C with incident CAD in Low, Intermediate, and High PRS groups. In each group, HRs were calculated relative to the near optimal (< 130 mg/dL) reference LDL-C level in a Cox proportional-Hazards model adjusted for age, gender, genotyping array, the first 4 principal components of ancestry, townsend deprivation index, diabetes and smoking status, family history of heart disease, systolic blood pressure, Glycated haemoglobin, Triglycerides, Body Mass Index, C-reactive protein, and dietary intake goals.

**eFigure 4.**
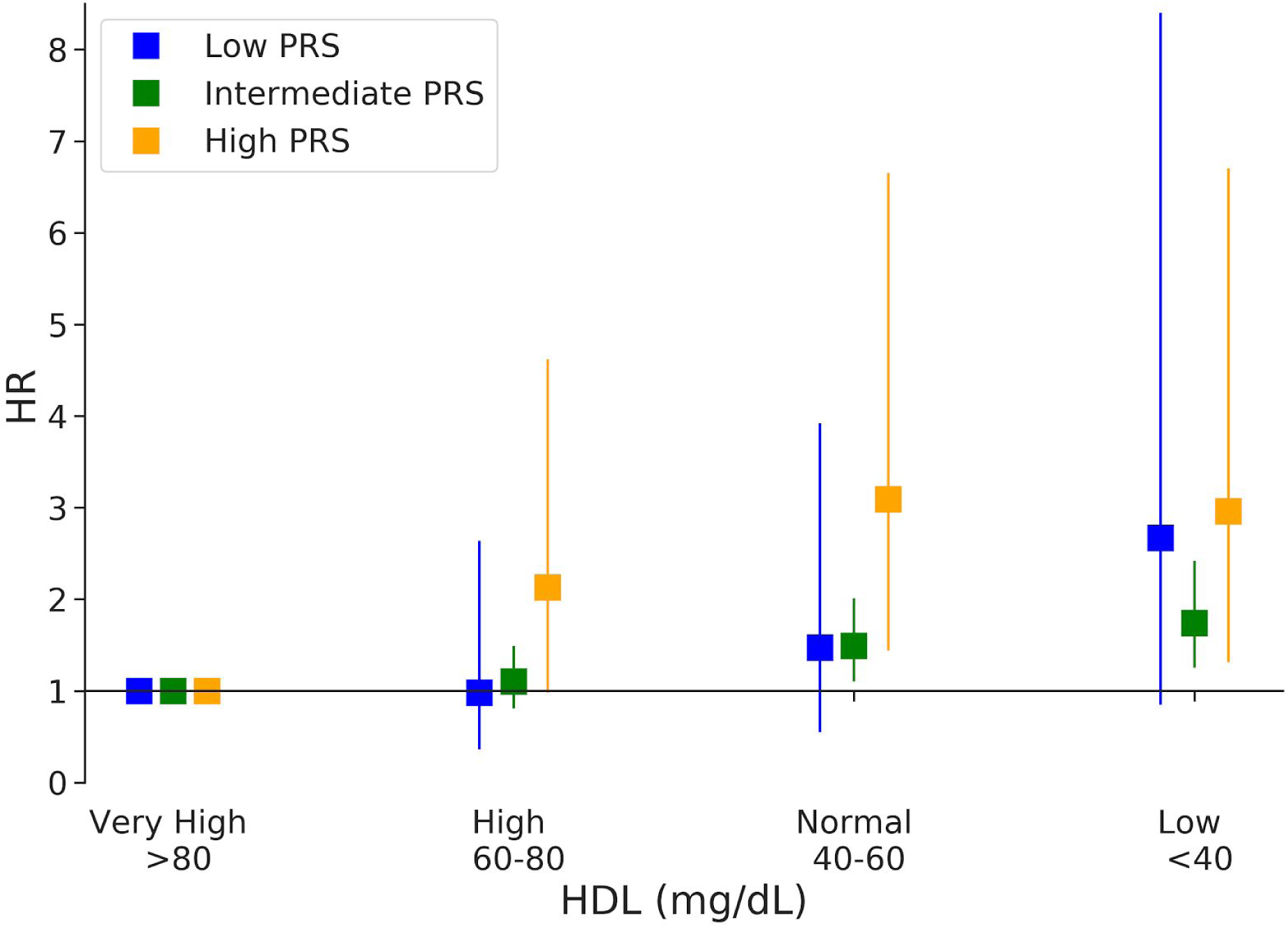
CAD risk by HDL in groups with different polygenic risk. Association of HDL with incident CAD in Low, Intermediate, and High PRS groups. HRs were calculated relative to the very high (> 80 mg/dL) reference HDL level in a Cox proportional-Hazards model adjusted for age, gender, genotyping array, the first 4 principal components of ancestry, townsend deprivation index, diabetes and smoking status, family history of heart disease, systolic blood pressure, Glycated haemoglobin, Triglycerides, Body Mass Index, C-reactive protein, and dietary intake goals.

